# Early effectiveness of COVID-19 vaccination with BNT162b2 mRNA vaccine and ChAdOx1 adenovirus vector vaccine on symptomatic disease, hospitalisations and mortality in older adults in England

**DOI:** 10.1101/2021.03.01.21252652

**Authors:** Jamie Lopez Bernal, Nick Andrews, Charlotte Gower, Julia Stowe, Chris Robertson, Elise Tessier, Ruth Simmons, Simon Cottrell, Richard Roberts, Mark O’Doherty, Kevin Brown, Claire Cameron, Diane Stockton, Jim McMenamin, Mary Ramsay

## Abstract

**Objectives:** To estimate the real-world effectiveness of the Pfizer/BioNTech BNT162b2 vaccine and Astrazeneca ChAdOx1 vaccine against Confirmed COVID-19, hospitalisations and deaths. To estimate effectiveness on the UK variant of concern.

**Design:** Test negative case control design

**Setting:** Community COVID-19 PCR testing in England

**Participants:** All adults in England aged 70 years and older (over 7.5 million). All COVID-19 testing in the community among eligible individuals who reported symptoms between 8^th^ December 2020 and 19^th^ February 2021 was included in the analysis.

**Interventions:** One and two doses of BNT162b2 vaccine. One dose of ChAdOx1 vaccine.

**Main outcome measures:** Symptomatic PCR confirmed SARS-CoV-2 infection, hospitalisations and deaths with COVID-19.

**Results:** Individuals aged >=80 years vaccinated with BNT162b2 prior to 4^th^ January, had a higher odds of testing positive in the first 9 days after vaccination (odds ratio up to 1.48, 95%CI 1.23-1.77), indicating that those initially targeted had a higher underlying risk of infection. Vaccine effectiveness was therefore estimated relative to the baseline post-vaccination period. Vaccine effects were noted from 10-13 days after vaccination, reaching an effectiveness of 70% (95% CI 59-78%) from 28-34 days, then plateauing. From 14 days after the second dose a vaccine effectiveness of 89% (95%CI: 85-93%) was seen.

Individuals aged >=70 years vaccinated from 4^th^ January had a similar underlying risk of COVID-19 to unvaccinated individuals. With BNT162b2, vaccine effectiveness reached 61% (95%CI 51-69%) from 28-34 days after vaccination then plateaued. With the ChAdOx1 vaccine, vaccine effects were seen from 14-20 days after vaccination reaching an effectiveness of 60% (95%CI 41-73%) from 28-34 days and further increasing to 73% (95%CI 27-90%) from day 35 onwards.

On top of the protection against symptomatic disease, cases who had been vaccinated with one dose of BNT162b2 had an additional 43% (95%CI 33-52%) lower risk of emergency hospitalisation and an additional 51% (95%CI 37-62%) lower risk of death. Cases who had been vaccinated with one dose of ChAdOx1 had an additional 37% (95% CI 3-59%) lower risk of emergency hospitalisation. There was insufficient follow-up to assess the effect of ChAdOx1 on mortality due to the later rollout of this vaccine. Combined with the effect against symptomatic disease, this indicates that a single dose of either vaccine is approximately 80% effective at preventing hospitalisation and a single dose of BNT162b2 is 85% effective at preventing death with COVID-19.

**Conclusion:** Vaccination with either a single dose of BNT162b2 or ChAdOx1 COVID-19 vaccination was associated with a significant reduction in symptomatic SARS-CoV2 positive cases in older adults with even greater protection against severe disease. Both vaccines show similar effects. Protection was maintained for the duration of follow-up (>6 weeks). A second dose of BNT162b2 provides further protection against symptomatic disease but second doses of ChAdOx1 have not yet been rolled out in England. There is a clear effect of the vaccines against the UK variant of concern.

## Introduction

On the 8^th^ December 2020 the UK became the first country to implement a COVID-19 vaccination programme following the approval of the Pfizer-BioNtech messenger RNA (mRNA) vaccine, BNT162b2, for emergency use.(1) The programme has since expanded to include the AstraZeneca adenovirus-vector vaccine, ChAdOx1 nCOV-19, and over 6.5 million individuals have now been vaccinated. The burden of COVID-19 in the UK remains high and early evidence on the effectiveness of vaccines is essential for informing policy decisions on the ongoing rollout of the programme and the use of other non-pharmaceutical interventions.(2)

During these first few weeks of the programme, the priority groups for vaccination included: (i) residents in a care home for older adults and their carers; and (ii) all those 80 years of age and over and frontline health and social care workers.(3) From the 18^th^ of January, rollout was expanded to all adults aged 70 years or older and those in clinically extremely vulnerable groups. Delivery was initially through hospital trusts and care homes, where possible, subsequently also through primary care providers and mass vaccination centres. Interim results from phase 3 clinical trials have found the BNT162b2 and ChAdOx1 vaccines to be highly effective using a two-dose schedule with a target interval of 3 weeks and 4 weeks respectively between doses.(4, 5) Data from the ChAdOx1 trial suggests that protection may be greater with a longer dosing interval.(5) A reanalysis of the BNT162b2 trial data suggests that a single dose of this vaccine has an efficacy of 92.6% in the early post-vaccination period.(6) Furthermore, with other vaccines an extended interval between the prime and booster doses typically provides a better immune response to the booster dose.(7, 8) Based on this evidence, the increasing incidence of COVID-19 in the UK and the need to rapidly vaccinate as many vulnerable people as possible, on the 20^th^ December 2020, the Joint Committee on Vaccination and Immunisation (JCVI) advised that the dose interval for both vaccines could be extended to up to 12 weeks. A policy decision was subsequently made to prioritise vaccinating as many people as possible with the first dose.

Also in December 2020, a new COVID-19 variant of concern (labelled VOC 202012/01) was found to be associated with increasing case numbers in Kent in South East England.(9) Recent analyses suggest that this variant has increased transmissibility and it has since become the dominant strain in large parts of the UK.(10, 11) The variant is characterised by 23 mutations, including mutations to genes encoding the spike protein, the target in the two vaccines currently in use, as well as the majority of vaccine candidates.(9) Concerns have been raised on the possible impact of the new variant on vaccine effectiveness.(12)

Public Health England have undertaken their first analysis of the early effect of COVID-19 vaccination using routine testing and vaccination data. The aims of this analysis were: (i) to estimate the effect of vaccination on confirmed COVID-19 in adults aged >=70 years with one and two doses; (ii) to estimate vaccine effectiveness against the new variant of concern, VOC 202012/01; (iii) to estimate case hospitalisation and case fatality rates among vaccinated and unvaccinated cases.

## Methods

This study estimates the effect of vaccination with the BNT162b2 and ChAdOx1 COVID-19 vaccines on laboratory confirmed symptomatic disease in individuals aged 70 years or older in England.

All individuals aged 70 years or older in England (over 7.5 million individuals) were eligible for inclusion.

A test negative case control design was used to estimate odds ratios for testing positive for SARS-CoV-2 in all vaccinated compared to unvaccinated individuals with compatible symptoms who were tested using PCR. Test negative case control designs are considered powerful study designs for estimating vaccine effectiveness and are used extensively for estimating effectiveness of influenza vaccines and vaccines against other respiratory viruses.(13–15) They have been found to have high concordance with randomised controlled trials.(16, 17) Vaccination status is compared in individuals testing positive for the target organism compared to those testing negative. Comparing to other individuals presenting for testing but testing negative helps to control for factors that are typically difficult to estimate in observational studies including differences in health seeking behaviour, access to testing and case ascertainment.

### Data sources

#### Outcome assessment

Testing for COVID-19 in the UK is conducted through hospital and public health laboratories for those with a clinical need as well as some healthcare workers (referred to as Pillar 1) and through community testing (referred to as Pillar 2).(18) Anybody can access a Pillar 2 test if they have coronavirus symptoms (high temperature, new continuous cough, loss or change in sense of smell or taste), or if they are part of a local or national mass testing programme. For this analysis, PCR testing data from Pillar 2 in individuals who reported having symptoms were included, data were extracted for all tests between 26^th^ of October 2020 and the 21^st^ of February 2021.

The mutations to the Spike (S) gene in VOC 202012/01 cause a reproducible S gene target failure (SGTF) in laboratories using a three target from Thermo Fisher (TaqPath) PCR assay.(9) Between week commencing 7^th^ of December 2020 and week commencing 25^th^ of January 2021, VOC 202012/01 accounted for between 98 and 100% of SGTF in England.(19) SGTF therefore provides a good proxy for identification of VOC 202012/01 without relying on sequencing. An analysis of the vaccine effects against COVID-19 detections with SGTF was undertaken restricted to data from laboratories using the TaqPath assay.

#### Exposure assessment

Testing data were linked to individual vaccination histories from the national vaccination register (the National Immunisation Management System, NIMS) using NHS number, date of birth, surname, forename and postcode. All COVID-19 vaccines administered in England are recorded in the NIMS by clinicians through point of care applications. The NIMS data were extracted on February 22^nd^ 2021 with immunisations to February 21^st^ 2021. To allow for delayed entry of data into NIMS only samples taken from February 19^th^ 2021 were included in analyses.

#### Secondary outcomes

The data were also linked to hospitalisation data from the Emergency Care Dataset (ECDS), which includes hospital admissions via emergency departments but not elective admissions, and to deaths data from NHS records.(20)

#### Covariates

There are a range of factors that may be associated with both the likelihood of being offered and/or accepting a vaccine and risk of exposure to SARS-CoV-2 or propensity to be tested. These include demographic factors, such as age, gender, index of multiple deprivation and ethnicity; geography and period (incidence of COVID-19 varied by region and by week over the study period, as did vaccine rollout); and care home status, care homes have been high exposure settings during the course of the pandemic.

Age, gender, DOB, ethnicity and address were extracted from the testing data and the NIMS. Addresses were used to determine index of multiple deprivation quintile and were also linked to Care Quality Commission (CQC) registered care homes using the Unique Property Reference Number (UPRN).(21) Data were restricted to those aged over 70 (defined as those aged 70 or older on 31^st^ of March 2021).

## Statistical methods

Logistic regression was used to estimate the odds of vaccination in PCR confirmed cases compared to those testing negative.

Only individuals swabbed within 0-10 days of symptom onset were included in the analysis because sensitivity of PCR testing decreases beyond 10 days after symptom onset.(22) Individuals only contribute their first positive test from 8^th^ December (as this was the date that the vaccination programme was introduced) and if they were not tested positive in the previous 6 weeks (which could have indicated a single prolonged illness episode). Individuals contributed a maximum of three randomly chosen negative results in the follow-up period after excluding any taken within three weeks before a positive test result, or after a positive result, which are more likely to be false negatives, or taken within <7 days of a previous negative sample, again, because these could represent a single illness episode. In addition, any negative tests with a symptoms date within 10 days after a previous symptom date were excluded for the same reason.

For the primary analysis those with a history of a previous positive PCR or antibody test at any time prior to 8^th^ December were excluded in order to estimate vaccine effectiveness in fully susceptible individuals.

The following possible confounders were included in the logistic regression model: age (in five-year age group, at 31 March 2021), gender, ethnicity, geography (NHS region), index of multiple deprivation, whether they were a care home resident and week of symptom onset.

In order to understand how quickly a vaccine effect takes to become apparent and when a full effect is first reached, as well as to better understand potential biases in the analysis, narrow follow-up windows are chosen (2 periods per week up to 14 days and weekly thereafter). Vaccination status was categorised by unvaccinated and the following time periods between vaccination and symptom onset: post dose 1: 0-3,4-6,7-9,10-13,14-20, 21-27, 28-34, 35-41, 42+; and post dose 2: 0-3, 4-6, 7-13, and 14+. Odd ratios were estimated for each period. For ChAdOx1 the final interval was 35+ due to the shorter follow-up time for this vaccine. For the analysis of either vaccine, individuals already vaccinated with the other vaccine were excluded.

Analyses were also stratified by vaccination period – prior to 4^th^ January (age 80+ only) and since the 4^th^ January (when the ChAdOx1 vaccine was introduced), SGTF (for vaccinations given in the period before Jan 4^th^, age 80+, BNT162b2 only). Comparison was to unvaccinated as the baseline group, however, for the earlier vaccination period and overall period a post-hoc analysis comparing to days 4-9 post-vaccination was also conducted to help account for the likely higher underlying risk of COVID-19 among those groups targeted for vaccination first.

The number of cases admitted to hospital within 14 days of the positive test, and the number of deaths within 21 days of the positive test, were estimated by vaccination status at the date of test (unvaccinated, vaccinated within 0-13 days, vaccinated at least 14 days before). Proportional hazards survival analyses were also conducted for these outcomes, adjusting for age, care home status, gender and period. This analysis was restricted to those over 80 years as this age group were targeted first and there is not yet enough follow-up in the 70-79 years age group to monitor these endpoints. To allow for delays in reporting of hospitalisations and deaths these data were censored at February 16^th^ and February 9^nd^, respectively for survival and 14 and 21 days earlier than this respectively for case-hospitalisation and case-fatality ratios. The analysis was repeated in test negative controls to assess healthy vaccine bias.

## Ethics

Surveillance of COVID-19 testing and vaccination is undertaken under Regulation 3 of The Health Service (Control of Patient Information) Regulations 2002 to collect confidential patient information (http://www.legislation.gov.uk/uksi/2002/1438/regulation/3/made) under Sections 3(i) (a) to (c), 3(i)(d) (i) and (ii) and 3(3). The study protocol was subject to an internal review by the PHE Research Ethics and Governance Group and was found to be fully compliant with all regulatory requirements. As no regulatory issues were identified, and ethical review is not a requirement for this type of work, it was decided that a full ethical review would not be necessary.

## Results

There were 174,731 pillar 2 PCR tested samples in individuals reporting symptoms within 10 days of the sample date. 156,930 of these (89.8%) successfully linked to vaccination data in the NIMS. 44,590 (28.4%) were positive tests, 112,340 (71.6%) were negative. The negative control samples were from a total of 108,851 individuals of whom 105,302 contributed one negative sample, 2,977 two samples, 256 three samples. 316 individuals contributed a negative sample and then a positive at least three weeks later. Differences in characteristics in linked and unlinked tests are shown in supplementary table 1. Characteristics were generally similar, though there was a higher proportion of non-white ethnic groups and individuals aged 85 years and older among those that did not link.

Vaccine coverage by vaccine brand at February 21^st^ 2021 within positive and negative tests is shown in table 1. This includes vaccines given both before and after onset date. It should be noted that vaccination with the BNT162b2 contributes more person-time due to the earlier rollout of this vaccine.

**Table 1:**
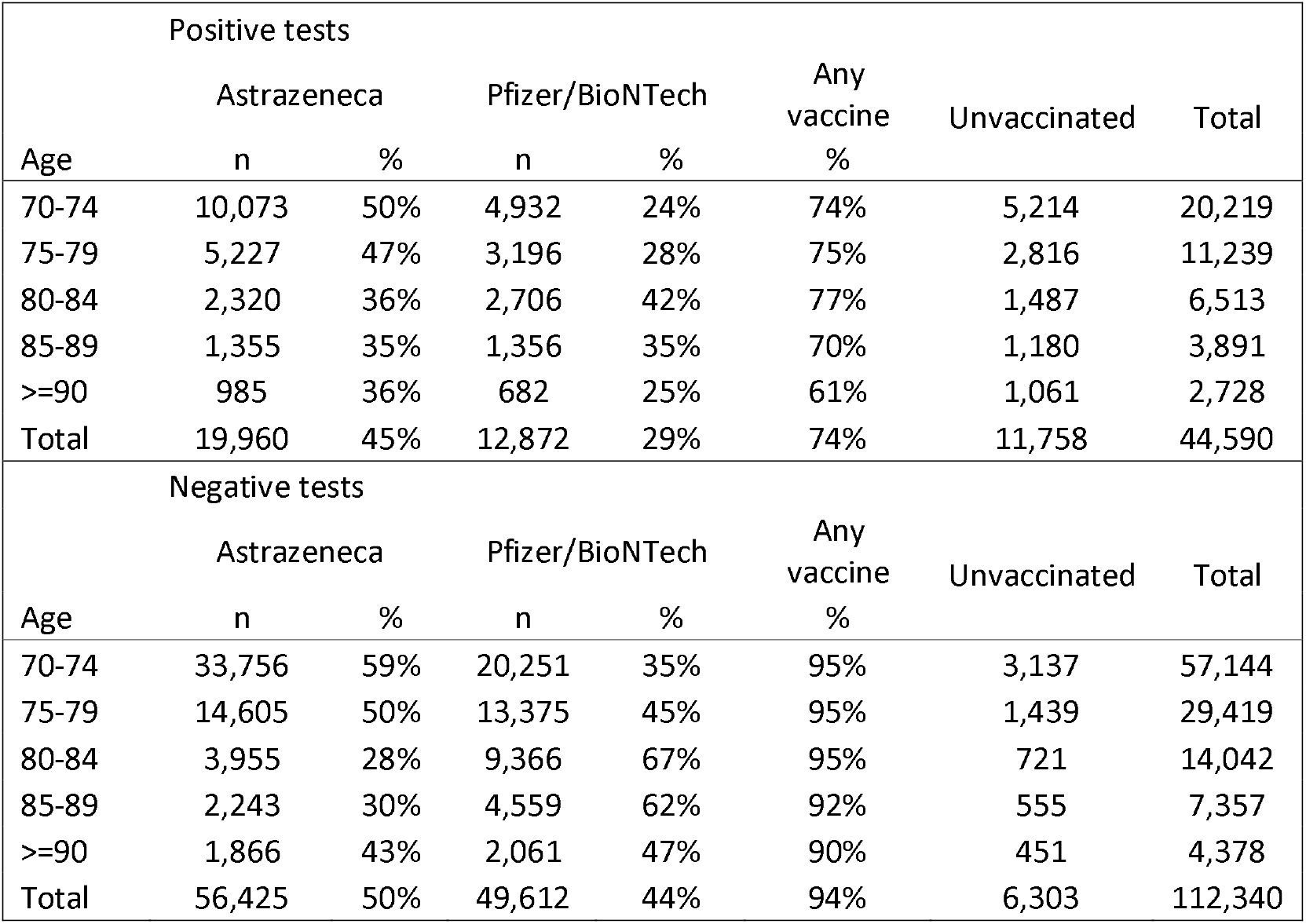
vaccine coverage by vaccine among cases and negative controls by the end of the study period (February 21^st^)by age group

Figure 1 shows the number of cases and controls by intervals around dose 1 and dose 2 of vaccination. The number of individuals tested beyond 42 days after vaccination with BNT162b2 is relatively small as are the numbers tested after 2 doses. The maximum follow-up after 1 dose was 56 days. With the ChAdOx1 vaccine the numbers tested beyond 28 days after vaccination were small with a maximum follow-up of 41 days. In the 7 days prior to vaccination the numbers of tests dropped and were mainly negative tests due to the requirement to defer vaccination recent COVID-19 cases. There was a notable increase in tests immediately after receiving the ChAdOx1 vaccine.

**Figure 1.**
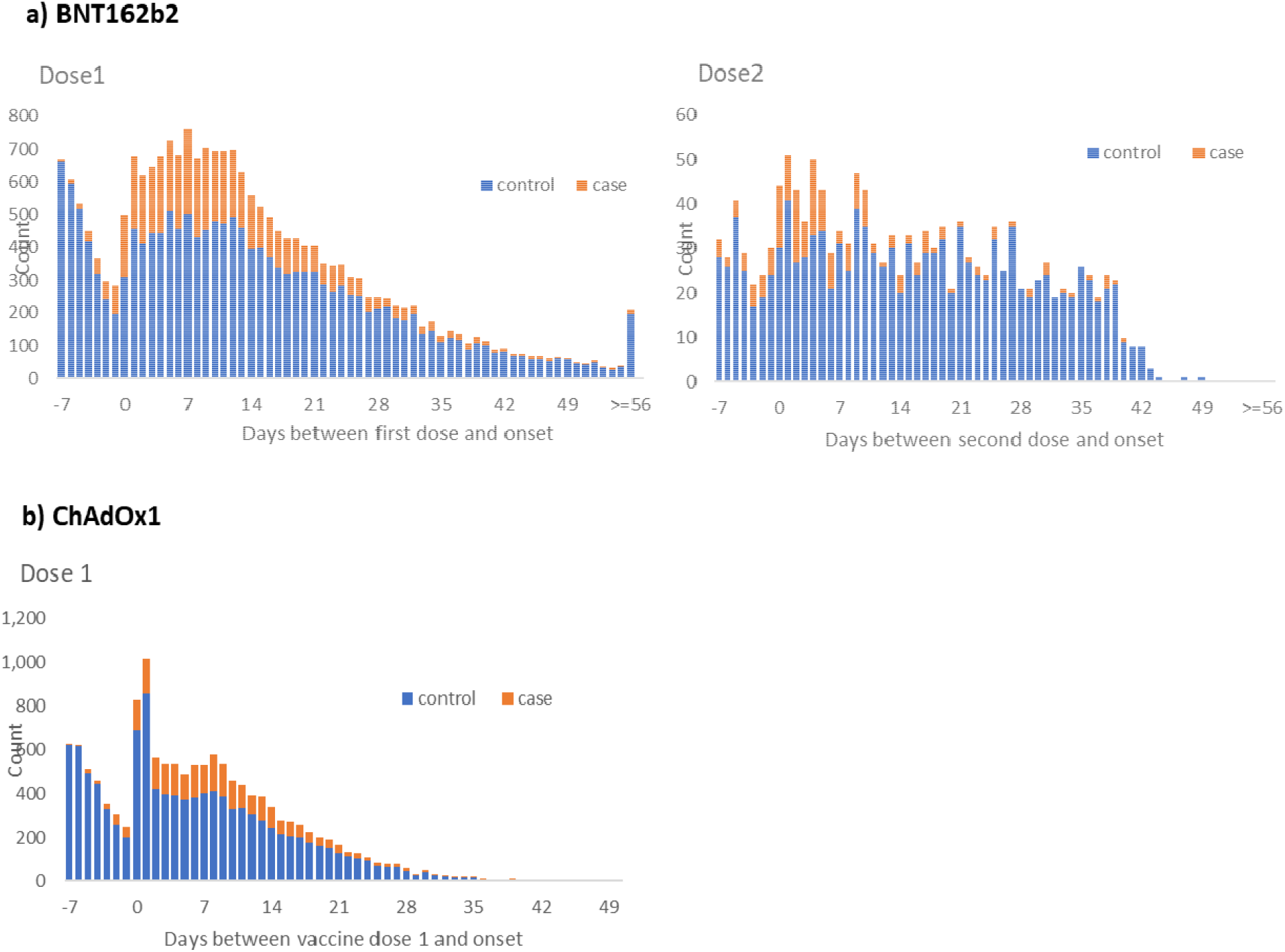
Cases and controls by interval from vaccination for BNT162b2 and ChAdOx1 vaccines

The odds of testing positive by interval after vaccination for BNT162b2 compared to those unvaccinated was initially analysed for the full period since the roll-out of the BNT162b2 vaccination programme on 8^th^ December 2020 (supplementary table 2, supplementary figure 1). During the first few days after vaccination (before an immune response would be anticipated), vaccinated individuals had a higher odds of testing positive, suggesting that vaccination was being targeted at those at higher risk of disease. The odds ratios then began to decrease from 14 days after vaccination, reaching 0.50 (95% CI 0.42-0.59) in the 28-34 day period and remaining stable thereafter. When those who had previously tested positive were included, results were almost identical (supplementary table 3). Stratifying by period indicated that vaccination prior to the 4^th^ of January was targeted at individuals at higher baseline risk of disease whereas since the 4^th^ of January (when the ChAdOx1 vaccine was introduced), delivery was more accessible for individuals with a similar baseline risk to the unvaccinated group. A stratified approach was therefore considered more appropriate for the primary analysis.

Results for BNT162b2 for vaccinations administered prior to the 4th of January are shown in Table 2 and Figure 2, this analysis was restricted to 80+ year olds as younger age groups were not eligible for vaccination prior to 4th of January. The odds of testing positive among vaccinated individuals increased during the early period up to days 7-9, reaching 1.48 (95%CI 1.23-1.77). The odds ratios then began to decrease from 10-13 days after vaccination, reaching 0.41 (95% CI 0.32-0.54) in the 28-34 day and remaining at a similar level from 35 days onwards. Compared to an unvaccinated baseline, this is equivalent to a vaccine effectiveness of 59%. Relative to the higher baseline risk seen during days 4-9, the odds ratio reaches 0.30 (95% CI 0.22-0.41) equivalent to a vaccine effectiveness of 70%. From 7 days after a second dose of BNT162b2 the odds ratio was 0.21 (95% CI 0.14-0.32) and then 0.15 (95% CI 0.11-0.21) from 14 days after the second dose, indicating vaccine effectiveness of 85%. Relative to the higher baseline risk seen during days 4-9, the odds ratio reaches 0.11 (95% CI 0.07-0.15) equivalent to a vaccine effectiveness of 89%.

**Table 2:** Adjusted odds ratios for confirmed case by interval after vaccination for BNT162b2, vaccinations administered prior to 4^th^ January 2021, age >=80 years

| Interval after dose<br>(days) |  | Vaccinated prior to 4th Jan |  |  |  |  |
| --- | --- | --- | --- | --- | --- | --- |
|  |  | controls | cases | OR (95% CI) | aOR (95% CI) | OR vs day 4-9 |
| unvaccinated |  | 15,718 | 8,988 | base | base |  |
| dose 1 | d1:0-3 | 277 | 167 | 1.17 (0.96-1.42) | 1.22 (1.00-1.48) |  |
|  | d1:4-6 | 241 | 179 | 1.26 (1.03-1.54) | 1.28 (1.05-1.56) |  |
|  | d1:7-9 | 252 | 257 | 1.47 (1.23-1.76) | 1.48 (1.23-1.77) |  |
|  | d1:10-13 | 361 | 284 | 1.12 (0.95-1.31) | 1.13 (0.96-1.33) | 0.82 (0.67-1.01) |
|  | d1:14-20 | 462 | 336 | 1.03 (0.89-1.19) | 1.06 (0.92-1.23) | 0.77 (0.63-0.94) |
|  | d1:21-27 | 288 | 118 | 0.60 (0.48-0.75) | 0.64 (0.51-0.79) | 0.46 (0.35-0.60) |
|  | d1:28-34 | 290 | 72 | 0.40 (0.30-0.52) | 0.41 (0.32-0.54) | 0.30 (0.22-0.41) |
|  | d1:35-41 | 274 | 65 | 0.45 (0.34-0.60) | 0.49 (0.37-0.66) | 0.36 (0.26-0.49) |
|  | d1:42+ | 396 | 59 | 0.34 (0.25-0.47) | 0.39 (0.29-0.55) | 0.28 (0.20-0.40) |
| dose 2 | d2:0-3 | 116 | 45 | 0.55 (0.39-0.77) | 0.59 (0.41-0.83) | 0.42 (0.29-0.62) |
|  | d2:4-6 | 80 | 30 | 0.52 (0.34-0.80) | 0.57 (0.37-0.88) | 0.41 (0.26-0.65) |
|  | d2:7-13 | 201 | 28 | 0.20 (0.13-0.29) | 0.21 (0.14-0.32) | 0.15 (0.10-0.23) |
|  | d2:14+ | 634 | 41 | 0.13 (0.09-0.18) | 0.15 (0.11-0.21) | 0.11 (0.07-0.15) |

d1= interval after dose 1, d2= interval after dose 2. OR: odds ratios period adjusted by week of onset. aOR: odds ratios adjusted for age, period, sex, region, ethnicity, care home, imd quintile

**Figure 2.**
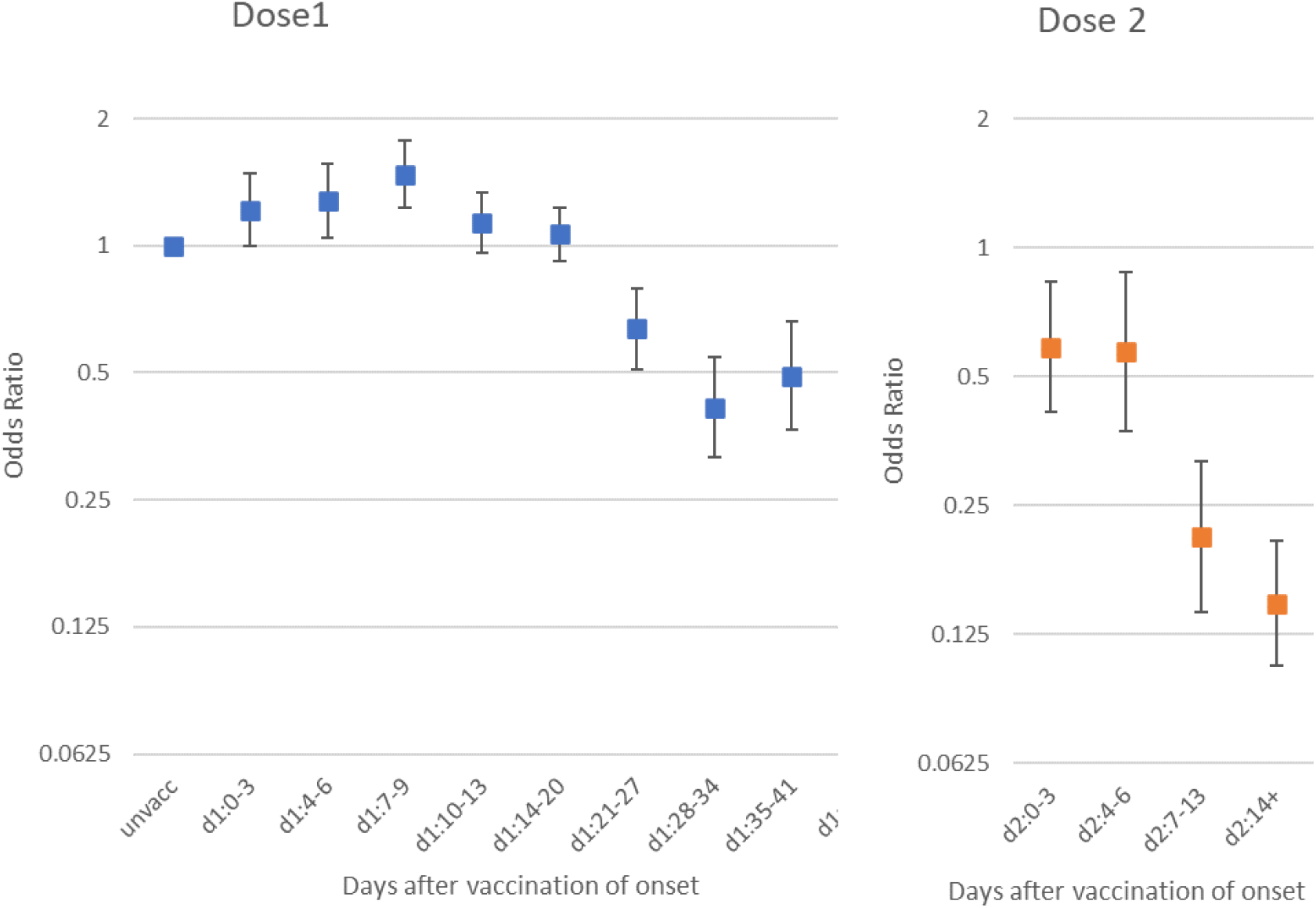
Adjusted odds ratios for confirmed case by interval after vaccination for BNT162b2, vaccinations administered prior to 4^th^ January 2021, age >=80 years

Results for both BNT162b2 and the ChAdOx1 vaccine administered since the 4th of January are shown in Table 3 and Figure 3. In this analysis, there was no significantly increased risk during the early post-vaccination period for either vaccine. For the ChAdOx1 vaccine, the odds ratio decreased during the 0-3 day period, which is associated with the increased testing immediately after vaccination noted above, and likely due to a reactogenicity effect of the vaccine. With BNT162b2, the odds ratio starts to decline from 10-13 days after vaccination, reaching 0.39 (95%CI 0.31-0.49) from 28 days after vaccination, equivalent to a vaccine effectiveness of 61% then remains at a similar level. With the ChAdOx1 vaccine the decline begins from 14-20 days after vaccination and reaches and odds ratio of 0.40 (95%CI 0.27-0.59) from 28 days after vaccination equivalent to a vaccine effectiveness of 60% and then reaches 0.27 (95%CI 0.10-0.73) from 35 days after vaccination, equivalent to vaccine effectiveness of 73% with wide confidence intervals. Confidence intervals for the two vaccines overlap and further follow-up is needed to understand whether the effects have plateaued for ChAdOx1. There were notable differences between the adjusted and unadjusted odds ratios in this analysis, but this was not seen in the pre-January the 4th analysis. This was due to confounding by age and care home status, likely because few care home residents were vaccinated in the early period and this period was restricted to a smaller age group (>=80 years). Results for the analysis of the ChAdOx1 vaccine including previous positives are shown in supplementary table 4 and show similar effects.

**Table 3:**
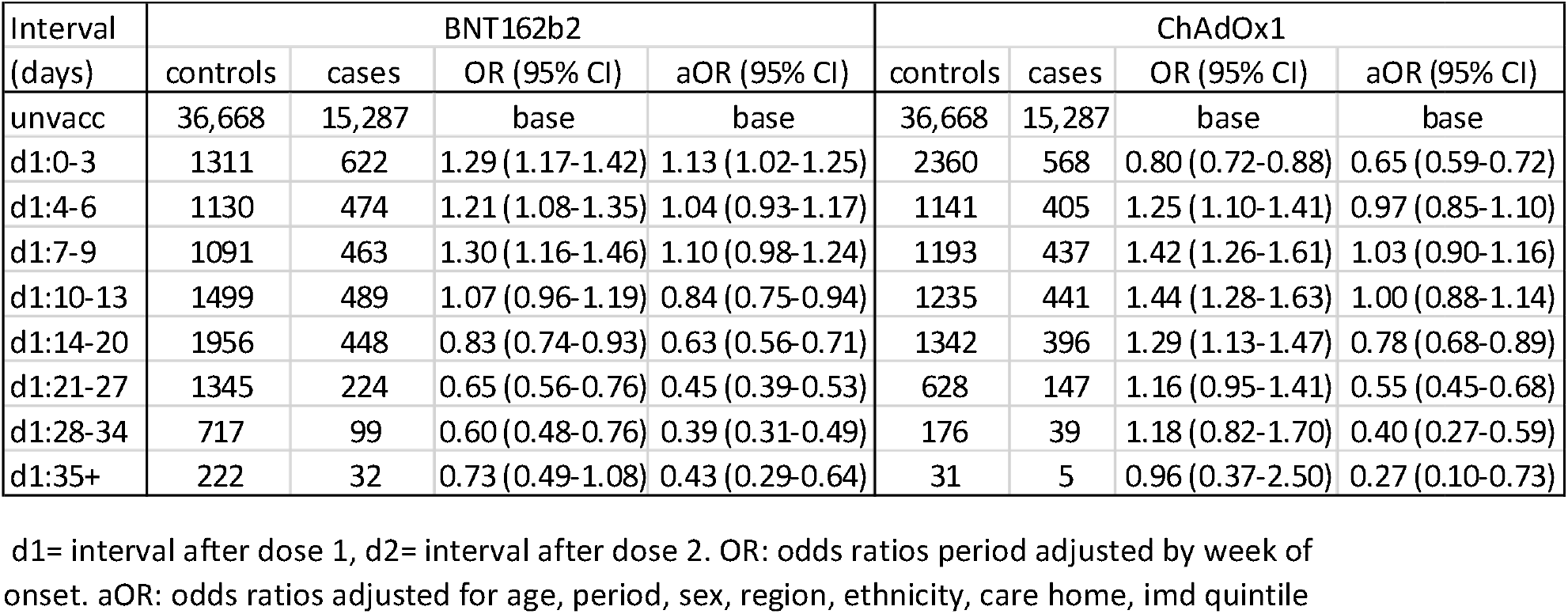
Adjusted odds ratios for confirmed case by interval after vaccination for both vaccines, vaccinations administered since 4^th^ January 2021, age >=70 years

| Interval<br>(days) | BNT162b2 |  |  |  | ChAdOx1 |  |  |  |
| --- | --- | --- | --- | --- | --- | --- | --- | --- |
|  | controls | cases | OR (95% CI) | aOR (95% CI) | controls | cases | OR (95% CI) | aOR (95% CI) |
| unvacc | 36,668 | 15,287 | base | base | 36,668 | 15,287 | base | base |
| d1:0-3 | 1311 | 622 | 1.29 (1.17-1.42) | 1.13 (1.02-1.25) | 2360 | 568 | 0.80 (0.72-0.88) | 0.65 (0.59-0.72) |
| d1:4-6 | 1130 | 474 | 1.21 (1.08-1.35) | 1.04 (0.93-1.17) | 1141 | 405 | 1.25 (1.10-1.41) | 0.97 (0.85-1.10) |
| d1:7-9 | 1091 | 463 | 1.30 (1.16-1.46) | 1.10 (0.98-1.24) | 1193 | 437 | 1.42 (1.26-1.61) | 1.03 (0.90-1.16) |
| d1:10-13 | 1499 | 489 | 1.07 (0.96-1.19) | 0.84 (0.75-0.94) | 1235 | 441 | 1.44 (1.28-1.63) | 1.00 (0.88-1.14) |
| d1:14-20 | 1956 | 448 | 0.83 (0.74-0.93) | 0.63 (0.56-0.71) | 1342 | 396 | 1.29 (1.13-1.47) | 0.78 (0.68-0.89) |
| d1:21-27 | 1345 | 224 | 0.65 (0.56-0.76) | 0.45 (0.39-0.53) | 628 | 147 | 1.16 (0.95-1.41) | 0.55 (0.45-0.68) |
| d1:28-34 | 717 | 99 | 0.60 (0.48-0.76) | 0.39 (0.31-0.49) | 176 | 39 | 1.18 (0.82-1.70) | 0.40 (0.27-0.59) |
| d1:35+ | 222 | 32 | 0.73 (0.49-1.08) | 0.43 (0.29-0.64) | 31 | 5 | 0.96 (0.37-2.50) | 0.27 (0.10-0.73) |

d1= interval after dose 1, d2= interval after dose 2. OR: odds ratios period adjusted by week of onset. aOR: odds ratios adjusted for age, period, sex, region, ethnicity, care home, imd quintile

**Figure 3.**
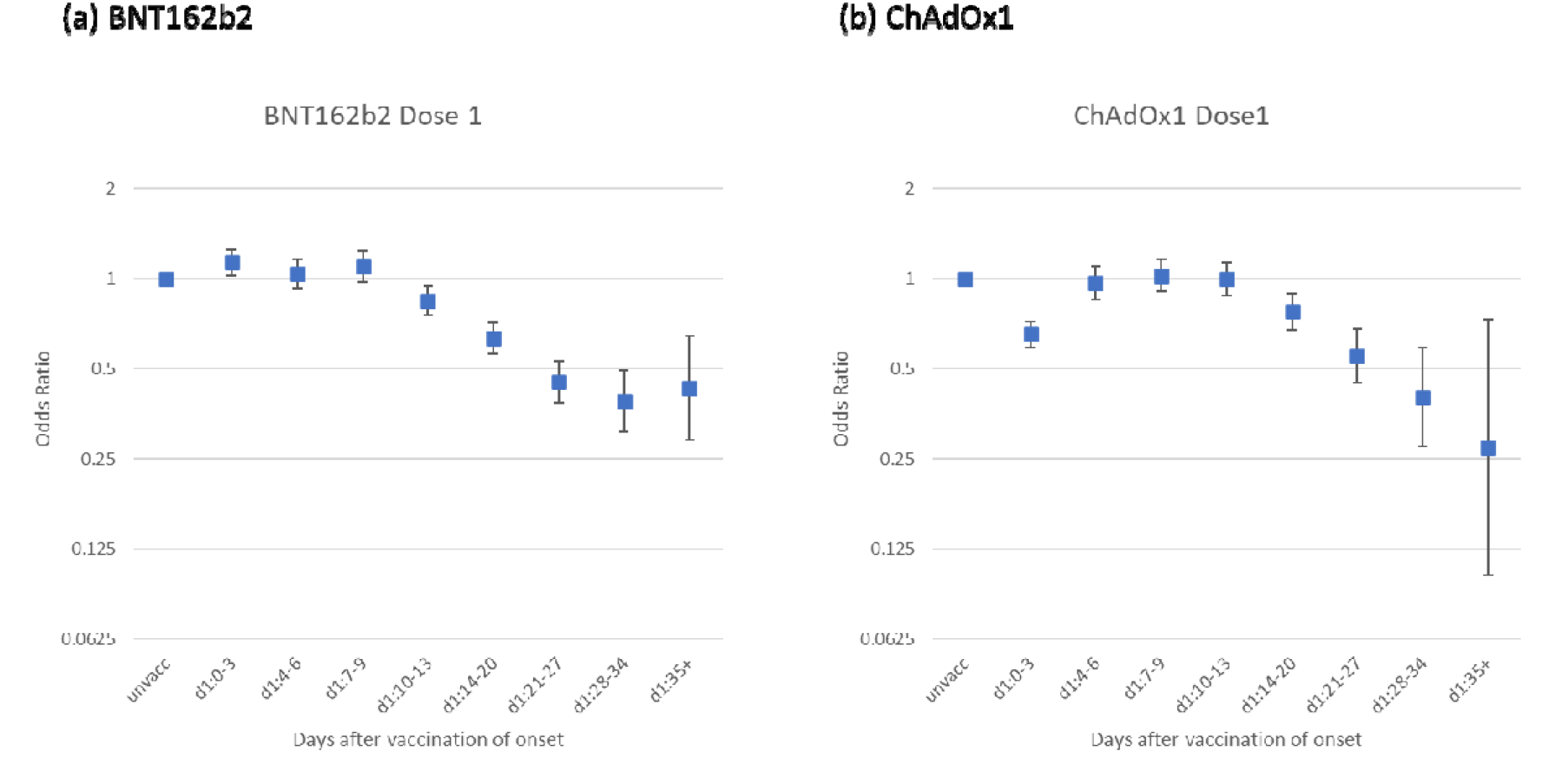
Adjusted odds ratios for confirmed case by interval after vaccination for BNT162b2 and ChAdOx1 vaccines, vaccinations administered since 4th January 2021

A further analysis by SGTF status to indicate those with and without VOC 202012/01 is shown in supplementary table 2. When using the comparison to the 4-9 day period to account for differences in baseline risk in those vaccinated, the results are similar with and without SGTF. There is a slightly bigger point estimate for vaccine effectiveness without SGTF in the 28-41 day periods but the effects are almost the same in the 42+ days period and confidence intervals overlap throughout. Numbers without SGTF are small, in particular during the later follow-up periods as VOC 202012/01 now dominates in England.

Table 5 shows hospital admissions in cases within 14 days of a positive test and deaths within 21 days of a positive test by vaccination status among those aged >=80 years. Hazard ratios from the survival analyses are also shown and the Kaplan-Meier curves are shown in supplementary figure 2. Hazard ratios for both vaccines were similar: 0.57 (95%CI: 0.48-0.67) and 0.63 (95%CI: 0.41-0.97) respectively for BNT162b2 and ChAdOx1 among those vaccinated at least 14 days prior to the test date, indicating that vaccinated individuals who do become symptomatic cases have an additional 43% and 37% protection against hospitalisation within 14 days of a positive test. The control analysis among those testing negative showed no difference in mortality rates by vaccination status indicating that there was no evidence of a healthy vaccinee effect (supplementary figure 4).

**Table 4:** Risk of admission to hospital within 14 days of testing positive in vaccinated and unvaccinated cases aged over 80 years

| Vaccination status | BNT162b2 |  |  |  | ChAdOx1 |  |  |  |
| --- | --- | --- | --- | --- | --- | --- | --- | --- |
|  | Total cases | Hospitalisations<br>n | Hospitalisations<br>% | Hazard ratio | Total cases | Hospitalisations<br>n | Hospitalisations<br>% | Hazard ratio |
| Unvaccinated | 8892 | 1365 | 15.35% | 1.00 | 8892 | 1365 | 15.35% | 1.00 |
| Test date less than 14 days after first dose | 2084 | 293 | 14.06% | 0.98 (0.86-1.11) | 562 | 64 | 11.39% | 0.98 (0.78-1.24) |
| Test date 14 days or more after first dose | 1400 | 128 | 9.14% | 0.57 (0.48-0.67) | 126 | 9 | 7.14% | 0.63 (0.41-0.97) |
| Total | 12376 | 1786 | 14.43% |  | 9580 | 1438 | 15.01% |  |

**Table 5:** Risk of death within 21 days of testing positive in vaccinated and unvaccinated cases aged over 80 years, BNT162b2

| Vaccination status | Deaths within 21 days |  |  |  |
| --- | --- | --- | --- | --- |
|  | Total cases | Deaths n | % | Hazard ratio |
| Unvaccinated | 8096 | 1063 | 13.13% | 1.00 |
| Test date less than 14 days after first dose | 1096 | 114 | 10.40% | 0.74 (0.62-0.90) |
| Test date 14 days or more after first dose | 750 | 51 | 6.80% | 0.49 (0.38-0.63) |
| Total | 9942 | 1228 | 12.35% |  |

Table 5 shows deaths within 21 days of a positive test by vaccination status among those aged >=80 years for BNT162b2. The hazard ratios for death compared to unvaccinated was 0.49 (0.38-0.63) for those vaccinated at least 14 days prior to the test date. This indicates that vaccinated individuals who do become symptomatic cases have an additional 51% protection against death within 21 days of a positive test. The control analysis among those testing negative showed no difference in mortality rates by vaccination status indicating that there was no significance evidence of a healthy vaccinee effect (supplementary figure 5).

## Discussion

This study provides the first real world evidence of COVID-19 vaccine effectiveness against symptomatic COVID-19 in older people in the UK. We find that a single dose of the BNT162b2 vaccine is approximately 60-70% effective at preventing symptomatic disease in adults aged 70 years and older in England and 2 doses are approximately 85-90% effective. Those vaccinated who went on to become a symptomatic case had a 44% lower risk of hospitalisation and a 51% lower risk of death compared to unvaccinated cases. We also provide the first real world evidence of effectiveness of the ChAdOx1 vaccine. The effect of a single dose of the ChAdOx1 vaccine against symptomatic disease was approximately 60-75% and there was again an additional protective effect against hospitalisation, though it is too early to assess the effect and mortality. VOC 202012/01 now dominates in the UK and these results will largely reflect vaccine effectiveness against this variant.

These data are observational and there are range of factors that influence the odds of testing positive which may also be associated with vaccination, thereby acting as confounders when examining vaccine effectiveness through routine testing, in particular in the early stages of the vaccination programme. A key factor that is likely to increase the odds of testing positive in vaccinees (therefore underestimating vaccine effectiveness) is that individuals initially targeted for vaccination may be at increased risk of exposure to COVID-19. For example, those accessing hospital may have been offered vaccine early in hospital hubs but may also be at higher risk of COVID-19. This may explain the higher odds of testing positive in vaccinees in the first few days after vaccination with the BNT162b2 (before they would have been expected to develop an immune response to the vaccine) among those vaccinated during the first month of the roll-out.(4, 23) This effect appears to lessen as roll-out vaccination programme progresses suggesting that access to vaccine was initially focussed on those at higher risk, although this bias may still affect the longer follow-up periods (to which those vaccinated earliest will contribute) more than the earlier follow-up periods. This may also mean that we might expect to see lower odds ratios in the later periods over time (i.e. estimates of vaccine effectiveness may increase further). In the opposite direction, vaccinees may have a lower odds of testing positive in the first few days after vaccination because individuals are asked to defer vaccination if they are acutely unwell, have been exposed or have had a recent coronavirus test.(24) This explains the lower odds of testing positive in the week prior to vaccination and may also persist for some time after vaccination if there are inaccuracies in the recording of symptom onset date. Vaccination can also cause systemic reactions including fever and fatigue.(23, 24) This may prompt more testing for COVID-19 in the first few days after vaccination, which, if due to a vaccine reaction, will be negative. This is likely to explain the increased testing immediately after receiving the ChAdOx1 vaccine and leads to an artificially low vaccine effectiveness in that period.(25, 26)

There are similarities between our results and those seen in the phase 3 clinical trials.(4, 5, 27) With BNT162b2, as in the trial, we begin to see a decline in the odds of testing positive among vaccinees from 10-13 days after the first dose. The trial found an overall efficacy of 94.7% after the second dose in those aged >= 65 years. We estimate a vaccine effectiveness of 90% in individuals aged 80 years or older. In the trial the reported vaccine effectiveness after dose 1 to before dose 2 was 52.4% (95% CI: 29.5-68.4%). However, this included cases from the first 2 weeks after vaccination when we wouldn’t expect any effect. Reanalysing the trial data using only cases observed between days 15 and 21 after the first dose, efficacy against symptomatic COVID-19 is estimated at 92.6% (95%CI: 69.0–98.3).(6, 28) Our analysis using the observational data suggests vaccine effectiveness against symptomatic disease reaches approximately 70% from 28 days after the first dose in those aged >=70 years. Real world evidence of BNT162b2 has also started to emerge from Israel on the early effectiveness of a single dose: Chodick et al estimated a vaccine effectiveness of 52% during the first 24 days after vaccination, though a reanalysis of the same data by Hunter et al estimated that by day 24, vaccine effectiveness had reached 90%.(29, 30). Dagan et al found a vaccine effectiveness against symptomatic disease of 57% in the 14-20 day period and 66% in the 21-27 day period which is similar to our estimates. Amit et al estimated a vaccine effectiveness of 85% from 15-28 days after the first dose, though they also estimated a 45% reduction from days 1-14 which may indicate that those vaccinated had a lower baseline risk.(31) It is not clear whether this analysis is based on symptom onset date or test date. The differences between some of the Israeli results and those seen in England may be explained by differences in the populations analysed, the case definitions or the analytical approach. Routine community testing may be being undertaken on the basis of non-specific symptoms. Another possible explanation is differential effectiveness against different variants. In England, VOC 202012/01 was the dominant virus throughout the study period. However, our analysis by variant based on SGTF suggests that there is little difference in effects by variant. This is supported by recent evidence that sera from vaccinated individuals elicits equivalent neutralising titres to VOC 202012/01 and similar variants to that seen with previous strains.(32, 33)

We also provide evidence that a single dose provides additional protection against hospitalisation and death, with cases among vaccinated individuals having around half the risk of these severe outcomes compared to unvaccinated cases. Combining this with our minimum vaccine effectiveness against symptomatic disease estimate, would suggest that a single dose of BNT162b2 is around 80% effective at preventing hospitalisation and around 85% effective at preventing death with COVID-19.

We found that the ChAdOx1 vaccine reaches 75% effectiveness from 35 days after the first dose in those aged 70 years and older. This had not yet plateaued so we are not able to estimate the level of vaccine effectiveness that will be reached with this vaccine or the duration of this effect. As with BNT162b2 there was additional protection against hospitalisation suggesting vaccine effectiveness against hospitalisation of at least 80% following a single dose of ChAdOx1. The initial phase 3 trial found a two-dose efficacy against symptomatic disease of 70.4% (all ages).(5) Efficacy results for older adults have not been reported but immune response was similar.(26) An analysis by duration of interval between doses suggested that a longer duration provided increased protection.(27) The efficacy of a single dose was estimated at 76% with follow-up up to 90 days for all age groups which is in line with our findings.(27)

This study has number of strengths: we have a large sample size, including all community COVID-19 testing in England since the start of the vaccination programme, data on symptoms and onset date, detailed vaccine history and data on all prior testing. We provide evidence of effectiveness without restricting to the defined populations and storage, maintenance and cold chains that can be well controlled in trial conditions but may be more challenging in the real world. The large sample size allows us to look at very fine intervals after vaccination which helps to understand possible biases that need to be accounted for in this early phase of the programme. The large sample size allows us to estimate effects on severe outcomes, which may not be possible from the trials. The observational nature of this analysis means that there are limitations and the results should be interpreted with caution. Some of the key confounding issues have been outlined above but there are others. Factors that could increase the risk of COVID-19 in vaccinees (and therefore result in underestimation of vaccine effects) include: individuals may have more risky behaviours after vaccination if they believe they are protected; also, presenting for vaccination may be a risk factor in itself (for example travelling to a vaccination centre with a friend or relative). Conversely, individuals who have been self-isolating may defer vaccination and may also be at lower risk of infection, this could underestimate vaccine effectiveness in the short period after vaccination. Misclassification is also likely to be a factor in this study. Symptoms are self-reported and may not be specific for COVID-19 without clinician diagnosis. Furthermore, individuals may falsely report symptoms in order to be tested – which will both include asymptomatic individuals in the symptomatic analysis and meant that symptom onset dates are incorrect. Low sensitivity or specificity of PCR testing may also mean that cases and controls are misclassified. Failure to exclude those with past infection because of low testing rates in wave 1 is another possibility. Lags in vaccination data could also lead to misclassification, however, we excluded the most recent two days from the analysis and a review of the NIMS data showed that it is more than 90% complete beyond 2 days after vaccination (supplementary figure 6). Furthermore, these lags would only affect the very early post vaccination period, which is not of primary interest in this analysis. Any misclassification would attenuate vaccine effects. Furthermore, at this stage in the vaccination programme, the length of follow-up in this analysis is very limited. Further estimates in the coming weeks will include larger sample sizes and longer follow-up. We found a higher proportion of non-white ethnic groups and individuals aged 85 years and older among those that we were unable to link to vaccination histories, this could affect generalisability of the results, however, given the high linkage rates overall and the fact that the study covers the whole population of England, this is unlikely.

This study provides early evidence that the vaccine is having a significant effect on COVID-19 cases in England. We see a clear effect of the first dose of vaccine, supporting the decision to maximise the number of individuals vaccinated with a single dose, though we have limited evidence on the duration of this effect. There are still significant numbers of vaccinated individuals who go on to develop COVID-19 and our study indicates that vaccinated individuals must maintain other precautions, in particular during the first two to three weeks after vaccination. We also provide evidence that BNT162b2 is effective at preventing severe disease. Further evidence is needed on the duration of any effect and the effect against asymptomatic infection and transmission and the four UK nations will work closely to develop and share evidence on this as it becomes available. Nevertheless, the fact that the vaccine appears to be preventing symptomatic disease, including with the new variant of concern, is encouraging and this is likely to have a significant impact on case detections and severe outcomes at a population level.

#### What is already known on this topic

- Clinical trials and emerging real-world data have shown that BNT162b2 is effective at preventing symptomatic disease using a two dose, 3 weeks apart schedule.
- Clinical trials have shown that ChAdOx1 is effective at preventing symptomatic disease in adults, though evidence in older adults is limited.

#### What this study adds

- A single dose of either vaccine provides significant protection against COVID-19 and further protection against severe disease lasting at least 6 weeks, including against the UK variant of concern.
- BNT162b2 and ChAdOx1 offer similar levels of protection in older adults.

## Statements

JLB, NA, CG and MR designed the study and developed the protocol and analysis plan. NA, CG, JS, CR cleaned and analysed the data. JLB drafted the manuscript. All authors contributed to the study design, and revised the manuscript. The corresponding author attests that all listed authors meet authorship criteria and that no others meeting the criteria have been omitted.

All authors have completed the ICMJE uniform disclosure form and declare: no support from any organisation for the submitted work; no financial relationships with any organisations that might have an interest in the submitted work in the previous three years, no other relationships or activities that could appear to have influenced the submitted work.

It was not appropriate or possible to involve patients or the public in the design, or conduct, or reporting, or dissemination plans of our research.

## Data Availability

Individual level data is not publically available.
Aggregated data on cases and vaccination rates are available via the following links: https://www.gov.uk/government/statistics/national-flu-and-covid-19-surveillance-reports and https://www.gov.uk/government/publications/covid-19-vaccine-monitoring-reports

## Acknowledgements

We would like to acknowledge the Public Health England COVID-19 Data Science Team, NHS England, NHS Digital and NHS Test and Trace for their roles in developing and managing the COVID-19 testing and vaccination systems and data sets as well as reporting NHS vaccinators, NHS laboratories, PHE laboratories and lighthouse laboratories. We would also like to thank the Joint Committee of Vaccines and Immunisations and the UK COVID-19 Vaccine Effectiveness Working Group for their advice and feedback in developing this study.

## Notes

### Competing Interest Statement

The authors have declared no competing interest.

### Funding Statement

This study was funded by Public Health England

### Author Declarations

PHE Research Ethics and Governance Group Statement: Surveillance of COVID-19 testing and vaccination is undertaken under Regulation 3 of The Health Service (Control of Patient Information) Regulations 2002 to collect confidential patient information (http://www.legislation.gov.uk/uksi/2002/1438/regulation/3/made) under Sections 3(i) (a) to (c), 3(i)(d) (i) and (ii) and 3(3). The study protocol was subject to an internal review by the PHE Research Ethics and Governance Group and was found to be fully compliant with all regulatory requirements. As no regulatory issues were identified, and ethical review is not a requirement for this type of work, it was decided that a full ethical review would not be necessary.

