## supplementary figures for "Early effectiveness of COVID-19 vaccination with BNT162b2 mRNA vaccine and ChAdOx1 adenovirus vector vaccine on symptomatic disease, hospitalisations and mortality in older adults in England"

Supplementary fig 1: Adjusted odds ratios for confirmed case by interval after vaccination for BNT162b2, age >=70 years since 8th December

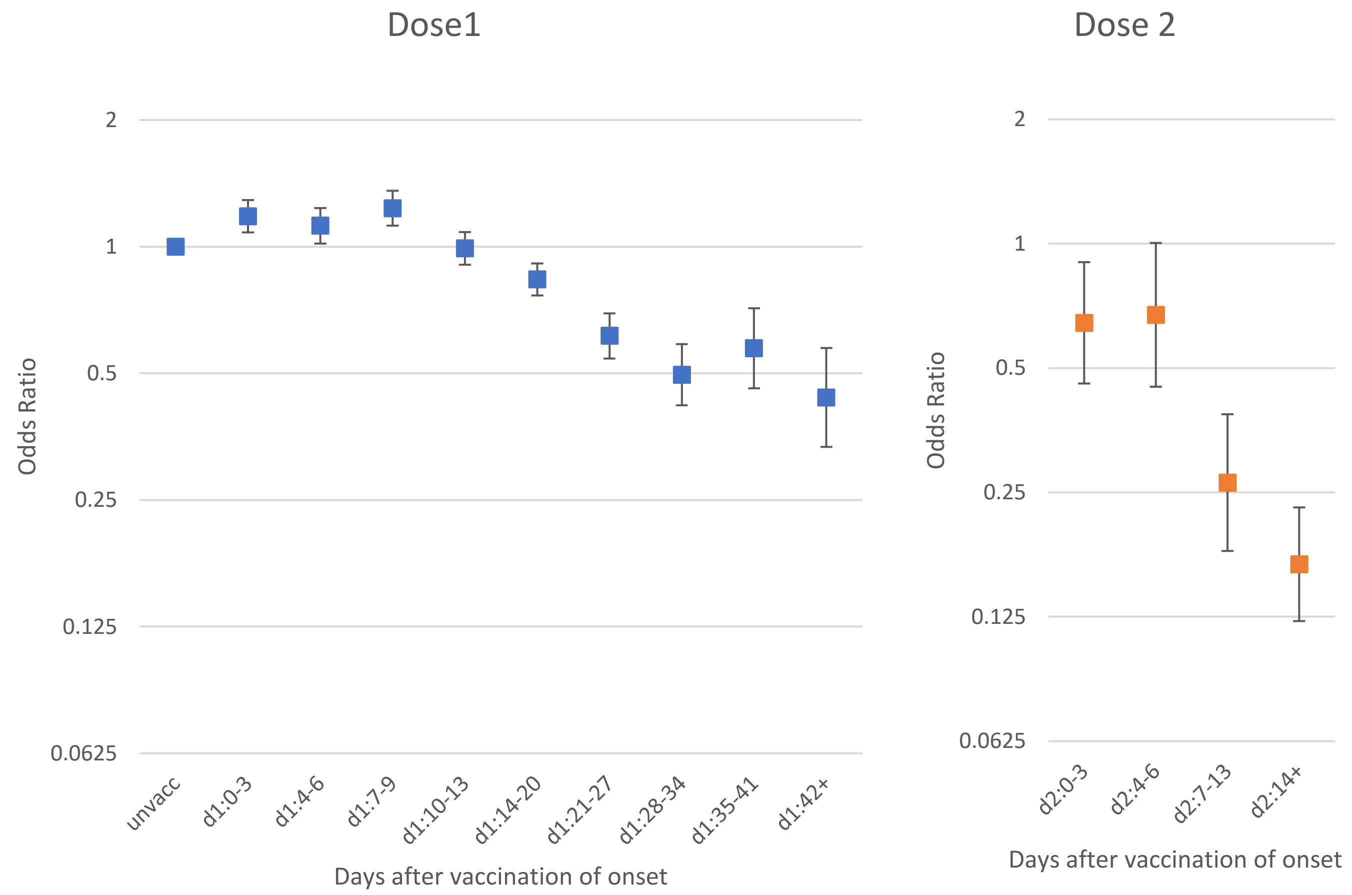

Supplementary fig 2: Kaplan-Meier curves for hospitalisations by vaccination status for BNT162b2 and ChAdOx1

(a) BNT162b2 (b) ChAdOx1

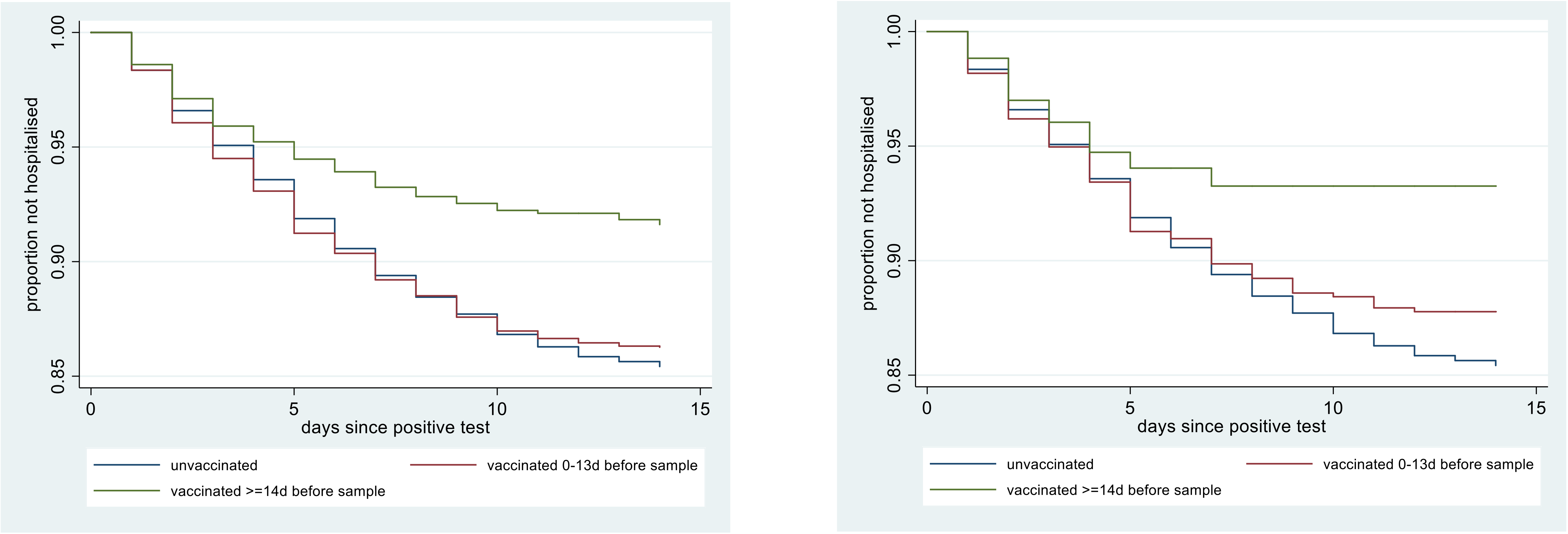


(a) BNT162b2 Adjusting for age, care home, period and gender the hazard ratios compared to unvaccinated were 0.98 (95% CI 0.86-1.11) for days 0-13 and 0.56 (0.47-0.67) for days 14+.

(b) ChAdOx1 Adjusting for age, care home, period and gender the hazard ratios compared to unvaccinated were 0.97 (95% CI 0.76-1.23) for days 0-13 and 0.58 (0.36-0.94) for days 14+.

Supplementary fig 3: Kaplan-Meier curves for deaths by vaccination status for BNT162b2

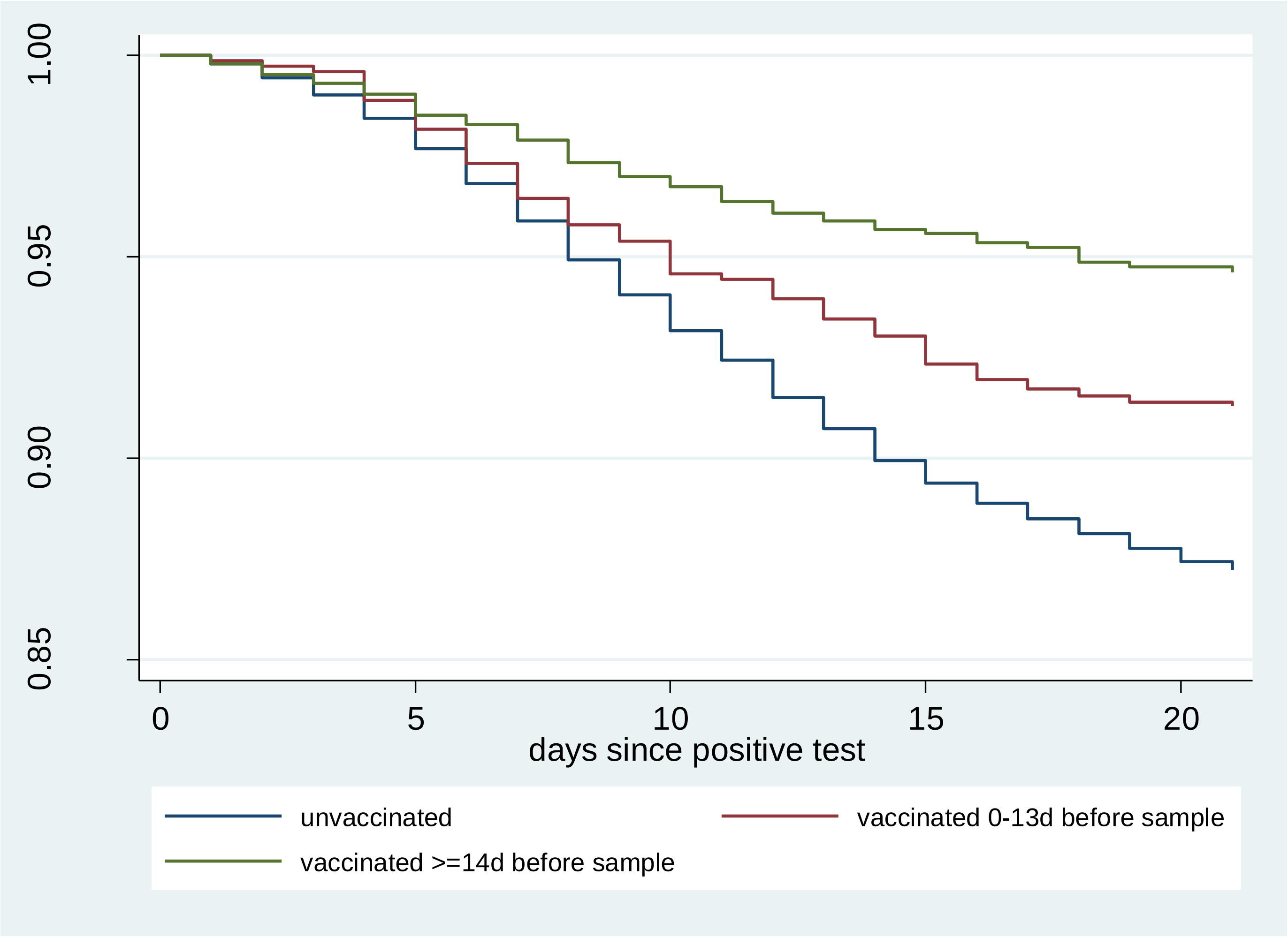


Adjusting for age, care home, period and gender the hazard ratios compared to unvaccinated were 0.74 (95% CI 0.62-0.90) for days 0-13 and 0.49 (0.38-0.63) for days 14+.

Supplementary fig 4: control analysis - Kaplan-Meier curves for hospitalisation among test negatives

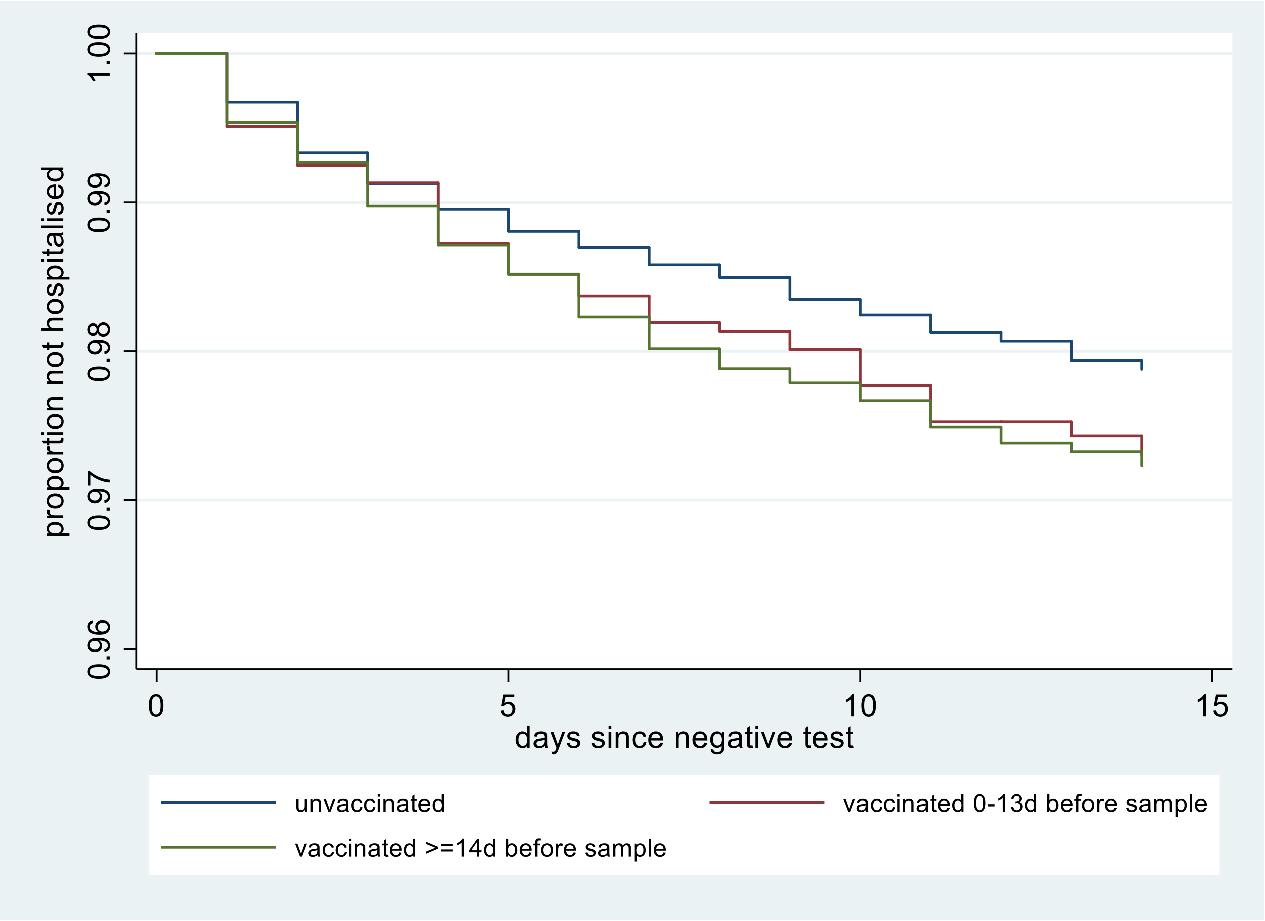


Adjusting for age, care home, period and gender the hazard ratios compared to unvaccinated were 1.24 (95% CI 0.98-1.56) for days 0-13 and 1.12 (0.89-1.41) for days 14+.

Supplementary fig 5: control analysis - Kaplan-Meier curves for deaths among test negatives BNT162b2

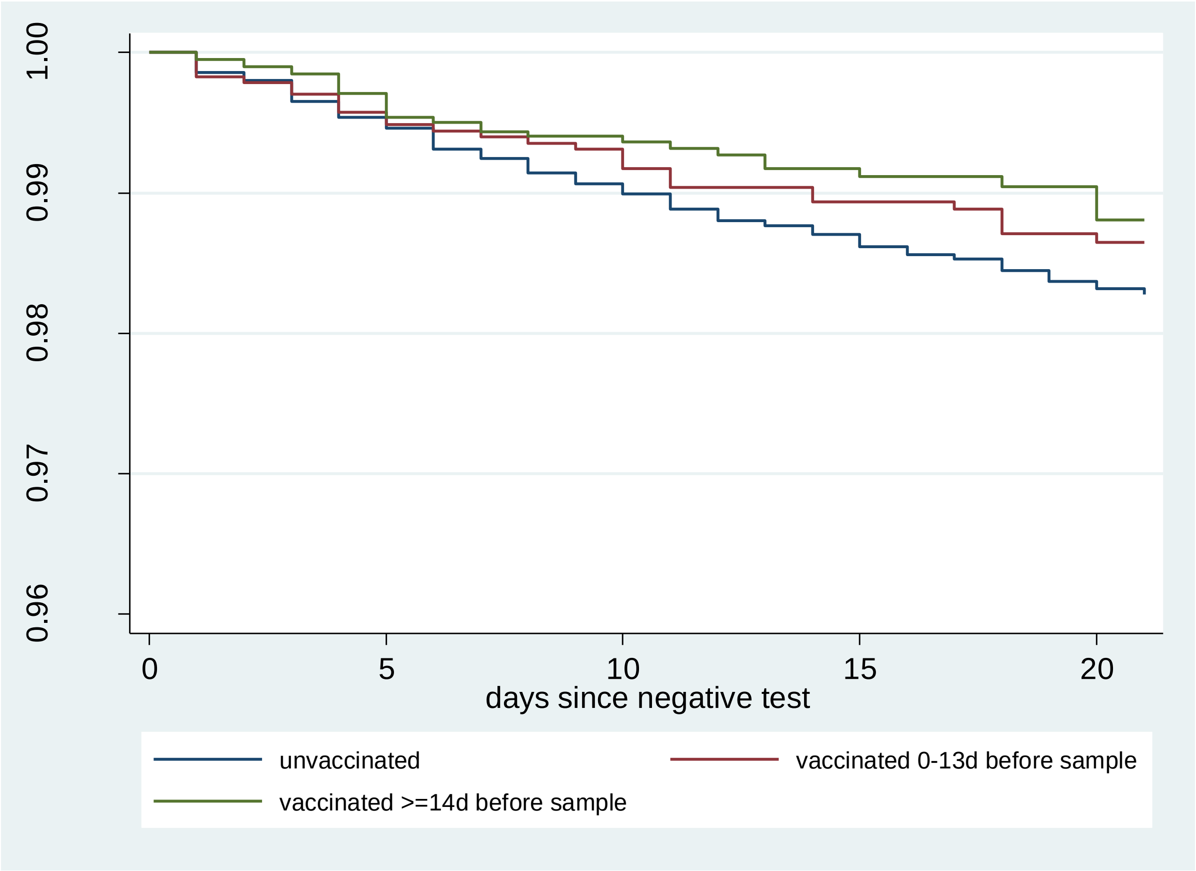


The Hazard ratios are 0.92 (0.62-1.35) and 0.81 (0.54-1.21) for intervals of 0-13 and 14+ from dose 1.

Supplementary fig 6 completeness of vaccination data by days after vaccination

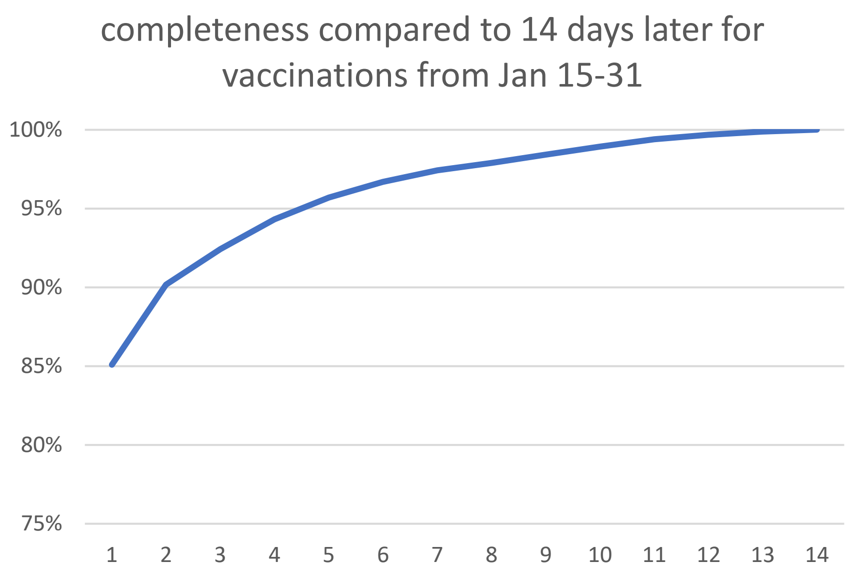
